# It’s risky to wander in September: modelling the epidemic potential of Rift Valley fever in a Sahelian setting

**DOI:** 10.1101/2020.02.25.20027821

**Authors:** Hélène Cecilia, Raphaëlle Métras, Assane Gueye Fall, Modou Moustapha Lo, Renaud Lancelot, Pauline Ezanno

## Abstract

Estimating the epidemic potential of vector-borne diseases, along with the relative contribution of underlying mechanisms, is crucial for animal and human health worldwide. In West African Sahel, several outbreaks of Rift Valley fever (RVF) have occurred over the last decades, but uncertainty remains about the conditions necessary to trigger these outbreaks. We use the basic reproduction number (*R*_0_) as a measure of RVF epidemic potential in Northern Senegal, and map its value in two distinct ecosystems, namely the Ferlo and the Senegal river delta and valley. We consider three consecutive rainy seasons (July-November 2014, 2015 and 2016) and account for several vector and animal species. Namely, we parametrize our model with estimates of *Aedes vexans arabiensis, Culex poicilipes, Culex tritaeniorhynchus*, cattle, sheep and goats abundances. The impact of RVF virus introduction is assessed every week, in 4367 pixels of 3,5km^2^. The results of our analysis indicate that September was the month with highest epidemic potential in each study area, while at-risk locations varied between seasons. We show that decreased vector densities do not highly reduce *R*_0_ and that cattle immunity has a greater impact on reducing transmission than small ruminants immunity. The host preferences of vectors and the temperature-dependent time interval between their blood meals are crucial parameters needing further biological investigations.

**Highlights:**

- September is a period of high Rift Valley fever epidemic potential in northern Senegal regardless of the year, but exact locations where epidemics might start change between rainy seasons.
- Decreased vector densities during the rainy season did not highly reduce the epidemic potential of at-risk locations.
- High levels of immunity in cattle populations reduce more Rift Valley fever virus transmission than a high immunity in small ruminants in our study area. This aspect should be investigated further for targeted vaccination campaigns.
- Precise estimates of vector feeding preferences and the temperature-dependent lenght of their gonotrophic cycle are key to ensure a good detection of at-risk pixels.

## Introduction

Vector-borne diseases (VBDs) represent a growing threat to animal and human health worldwide. They account for 17% of all infectious diseases, affecting more than one billion people each year (WHO, 2014). Their presence in livestock can dramatically impact food production locally and hamper exportations (Davies and Martin, 2003). This burden mostly affects low-income countries and their socio-economic development (WHO, 2014). In addition, climate change along with increased people and animal mobility create opportunities for vectors and their pathogens to establish in new areas, as was the case for West Nile virus in the United States (Calisher, 2000). Developping efficient countermeasures against VBDs requires a good understanding of their transmission dynamics. This remains a major challenge considering the complexity of the biological system formed by pathogens, hosts, vectors and their relation to the environment (Parham et al., 2015).

Rift Valley fever virus (RVFV, Bunyaviridae : *Phlebovirus*) is a zoonotic and vector-borne pathogen, present throughout Africa, in the Arabian Peninsula and in the South West Indian Ocean islands. Mosquitoes of the *Aedes* and *Culex* genus are the main vectors (Chevalier et al., 2010), some of which are suspected to transmit the virus transovarially (Linthicum et al., 1985). They transmit it to a variety of domestic host species, including cattle, goats, sheeps and camels, causing storms of abortions and a high mortality in young animals (Pepin et al., 2010). Human infection can occur through mucous membrane exposure or inhalation of viral particles (Davies and Martin, 2003). Most cases are limited to mild ‘flu-like’ symptoms (Laughlin et al., 1979), but severe forms of the disease can be fatal. The case fatality rate is usually below 1% (Madani et al., 2003) but tends to increase in recent outbreaks (Chevalier et al., 2010). Spillover into human population mainly concerns people working in close contact with animals such as pastoralists, butchers or veterinarians (Anyangu et al., 2010; Linthicum et al., 2016), but can be a concern for the general population, e.g. in a context of massive slaughters during religious festivals (EMPRES, 2003; Lancelot et al., 2019). Vector-to-human transmission is possible but does not seem to be the major route of infection (Gerdes, 2004). Animal-to-animal transmission by direct contact seems possible but is not yet confirmed (Chevalier et al., 2010). Since 2015, RVF is part of the R&D Blueprint programme of the World Health Organization (Mehand et al., 2018).

Models are a powerful tool to explore pathogen transmission dynamics, and several approaches have already been used to answer questions about RVF virus emergence and spread (Métras et al., 2011; Danzetta et al., 2016). Statistical models evidenced an association between El Niño events and RVF occurrence in the Horn of Africa for the 2007 epidemic in Kenya (Anyamba et al., 2009), as well as the link between rainfall patterns and population dynamics of RVF vectors (Mondet et al., 2005). Network models highlighted factors influencing host mobility in regions affected by RVF (Apolloni et al., 2018; Kim et al., 2018; Belkhiria et al., 2019). The use of remote-sensing and geographic information systems (GIS) enabled the identification of landscape properties associated with RVF virus transmission (Tourre et al., 2009; Tran et al., 2016). However, prior to studying the transmission dynamics of a pathogen at large time- and spatial-scale, it is critical to understand the local impact of its introduction and in particular, its potential to trigger the onset of an epidemic.

The use of compartmental models together with the next generation matrix provides a way to estimate the basic reproduction number *R*_0_ and gain a deeper understanding of the underlying processes contributing to the epidemic potential (Hartemink et al., 2008). *R*_0_ represents the average number of secondary cases produced by one infected individual introduced in an entirely susceptible population over the course of its infectious period. Several theoretical mechanistic models have been proposed to formulate *R*_0_ for RVF (Gaff et al., 2007; Niu et al., 2012; Xue and Scoglio, 2013; Pedro et al., 2016), without being applied to real areas. However, *R*_0_ is context-specific and studies mapping *R*_0_ in space using data on hosts and vectors were done only for RVF-free regions, such as the Netherlands (Fischer et al., 2013) or the United States (Barker et al., 2013). In regions with regular RVF outbreaks, such as the West African Sahel, modelling *R*_0_ could explain what locally drives the rapid increase in RVF incidence and creates amplification hotspots.

Senegal and Mauritania have experienced several outbreaks since 1987. Most cases were reported in the Sahel region, more specifically in Northern Senegal and Southern Mauritania (Caminade et al., 2014; Sow et al., 2016). This region encompassing semi-arid to arid climate bridges the gap between the Sahara desert and the tropical rainforests of equatorial Africa. In Northern Senegal, RVF outbreaks have mainly been reported in two distinct ecosystems, along the Senegal river and in the Ferlo region. The hypotheses underlying RVF epidemic potential are assumed to differ between these two study areas. Indeed, along the Senegal River, water, vectors (mainly *Culex*) and hosts are in contact year round due to the development of irrigated agriculture (Bruckmann, 2018). In contrast, the Ferlo is much dryer. When the rainy season starts in July, temporary ponds are flooded and *Aedes* eggs, layed at the edges of ponds the year before, hatch and induce a rapid and massive emergence of adult mosquitoes (Ndione et al., 2008). In the meantime, vegetation grows and creates the suitable conditions for nomadic herds to stop during their transhumance pathway (Adriansen, 2008). Therefore, the presence at the same place of mosquitoes and livestock, which could both introduce the virus, create opportunities for RVF outbreaks.

Previous studies mapping RVF risk in West African Sahel overlapped climate anomalies and host densities, but without linking mechanistically the underlying processes to the disease outcome (Caminade et al., 2011, 2014). At very local scales, in particular around the village of Barkedji in the Ferlo region of Senegal, different approaches such as remote-sensing (Lacaux et al., 2007; Ndione et al., 2009) or statistical models (Bicout and Sabatier, 2004; Vignolles et al., 2009; Talla et al., 2016) were used. These studies focused on the link between landscape features (typically ponds, Soti et al., 2013; Bop et al., 2014) and vector abundance. Therefore, there is still a need for an indicator integrating all the major mechanisms suspected to play a role in RVF epidemic potential.

The aim of the present paper is to map the epidemic potential of RVF virus in a Sahelian setting during the rainy season, comparing two different study areas, namely the Senegal river delta and valley, and the Ferlo. For this, we give an expression of *R*_0_ in a multi-species (2 hosts and 2 vectors) context, accounting for vector feeding preferences. We identify parameters varying in time and space as well as relevant data sources to map contact zones between hosts and vectors. Then, we map the local epidemic potential for three consecutive rainy seasons for weekly dates of virus introduction. We identify locations and introduction times with higher epidemic potential. We assess the role of vector densities and herd immunity to reduce *R*_0_. Eventually, we test the robustness of our results through a sensitivity analysis.

## Material and methods

We built a compartmental, mechanistic model of RVF virus (RVFV) transmission with 2 host and 2 vector populations (Figure 1, Eq. S.1-S.3). We only included mechanisms accurately occuring at the onset of a potential epidemic, locally, upon the introduction of the virus, either by a host or a vector. The model was used to obtain the next-generation matrix. We derived the expression of the basic reproduction number *R*_0_ by using the method by van den Driessche and Watmough 2002. The value of *R*_0_ was computed for all pixels (of resolution 3.5 km^2^) containing both hosts and vectors, and for weekly dates of virus introduction spanning three rainy seasons (July to November) of 2014, 2015, and 2016. Dates and location of RVFV introduction were assumed independent from each other. In addition, host infectious period is rather short (around a week) and temperatures did not strongly vary in our study area (Figure S.2) over the course of a vector lifetime (around a month). Thus, parameters were kept constant within each *R*_0_ computation (i.e. each date and location of introduction) for the whole duration of secondary case generation, but were updated at each new computation.

**Figure 1:**
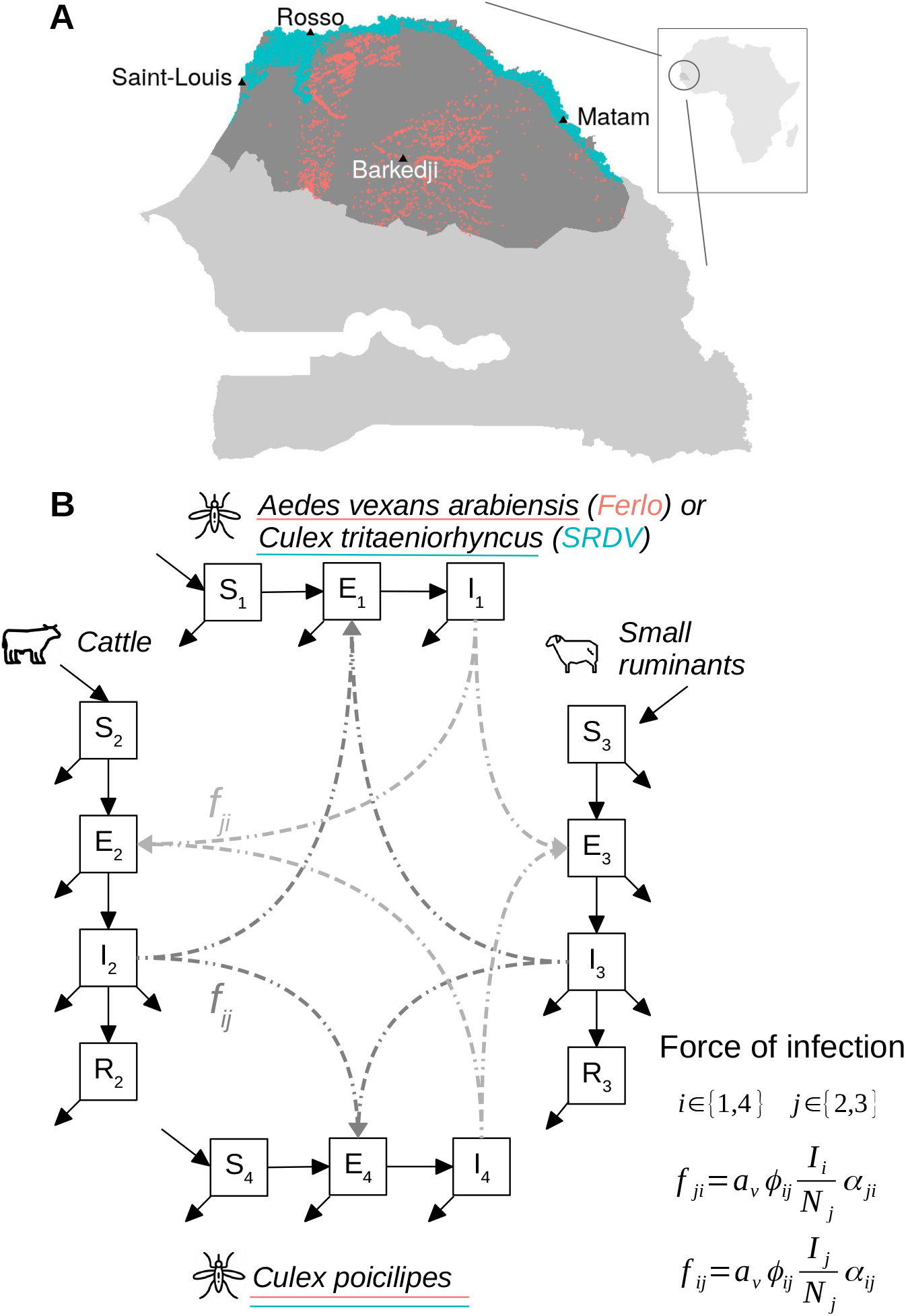
A - Study area, northern Senegal. Pixels highlighted correspond to locations with hosts and vectors at least once in the 3 rainy seasons, the colour indicating the region and thus the vector species they contain. Ferlo (pink), n = 1702, Senegal river delta and valley (SRDV, blue), n = 2665. B - Flow diagram of the RVFV mechanistic model used to obtain the next-generation matrix and derive the analytical formula of the basic reproduction number *R*_0_. Formulas give the force of infection in host populations (from vectors) *f*_*ji*_ (light grey) and in vector populations (from hosts) *f*_*ij*_ (dark grey).

### Model structure and assumptions

Vectors were modelled using susceptible (*S*_*i*_), latently infected (*E*_*i*_), and infectious (*I*_*i*_) health states, *i ∈ {*1, 4*}*. In the Ferlo, vector species considered were *Aedes vexans arabiensis* (subscript 1) and *Culex poicilipes* (subscript 4). In the Senegal river delta and valley (SRDV), vector species considered were *Culex tritaeniorhynchus* (subscript 1) and *C. poicilipes* (subscript 4). This was based on previous entomological studies conducted in both areas (Diallo et al., 2011; Fall et al., 2011; Biteye et al., 2018). *Ae. v. arabiensis* and *C. poicilipes* are confirmed vectors of RVFV in Senegal (Fontenille et al., 1998; Diallo et al., 2000; Ndiaye et al., 2016). *C. tritaeniorhynchus* is highly abundant and was identified as a RVFV vector in the 2000 outbreak in Saudi Arabia (Jupp et al., 2002). In the model, vectors were assumed to become infected either after biting infectious cattle or small ruminants, but could not transmit the virus transovarially. Whilst limited evidence of vertical transmission of RVFV in mosquitoes is available (Linthicum et al., 1985), we assumed that this mechanism would be related to interannually patterns, rather than epidemic potential during a given rainy season (Lumley et al., 2017).

Host populations contained susceptible (*S*_*j*_), latently infected (*E*_*j*_), infectious (*I*_*j*_), and recovered (with immunity, *R*_*j*_) individuals, *j ∈ {*2, 3*}*. They were stratified into cattle (subscript 2) and small ruminants (i.e goats and sheeps, subscript 3). Livestock could only be infected by the bites of infectious vectors. Animal-to-animal transmission by direct contact was here considered marginal compared to vector transmission, playing a minor role at the onset of a potential epidemic. Even though it might explain observed endemic patterns observed in unfavourable areas for mosquitoes (Nicolas et al., 2014), this transmission route has yet to be documented.

Mosquito biting rate, mortality and extrinsic incubation period (defined as the time between infection through a blood meal and virus presence in the salivary glands, Tjaden et al., 2013) were assumed to be temperature-dependent for all vector species. In addition, we assumed that a proportion *c*_*i*_ of mosquito populations *i* could have double, partial, blood meals (Table 1, Supplementary Information 1.2). Transmission was modelled as reservoir frequency-dependent (Figure 1, Eq. S.1), as defined by Wonham et al. (2006). This means that an individual vector was considered to have a constant contact rate (biting rate + feeding preferences) with livestock populations regardless of surrounding vector densities, whereas an individual host had a contact rate with vectors dependent on the vector-to-host ratio in the area (Gubbins et al., 2008). This type of transmission function can induce unreallistically high *R*_0_ values when livestock densities are too low or vector densities are too high (Wonham et al., 2006). Therefore, for each introduction date, we removed pixels with vector-to-host ratio (*N*_1_ + *N*_4_)*/*(*N*_2_ + *N*_3_) *>* 1000. The force of infection included the relative preference of vectors for both livestock populations (*π*_*ij*_, Table 1) combined with relative abundance of hosts to compute the proportion of blood meals taken on each host population (parameter *ϕ*_*ij*_, Table 1). Parameter values and references are in Table 1, Supplementary information 1.2.

**Table 1:**
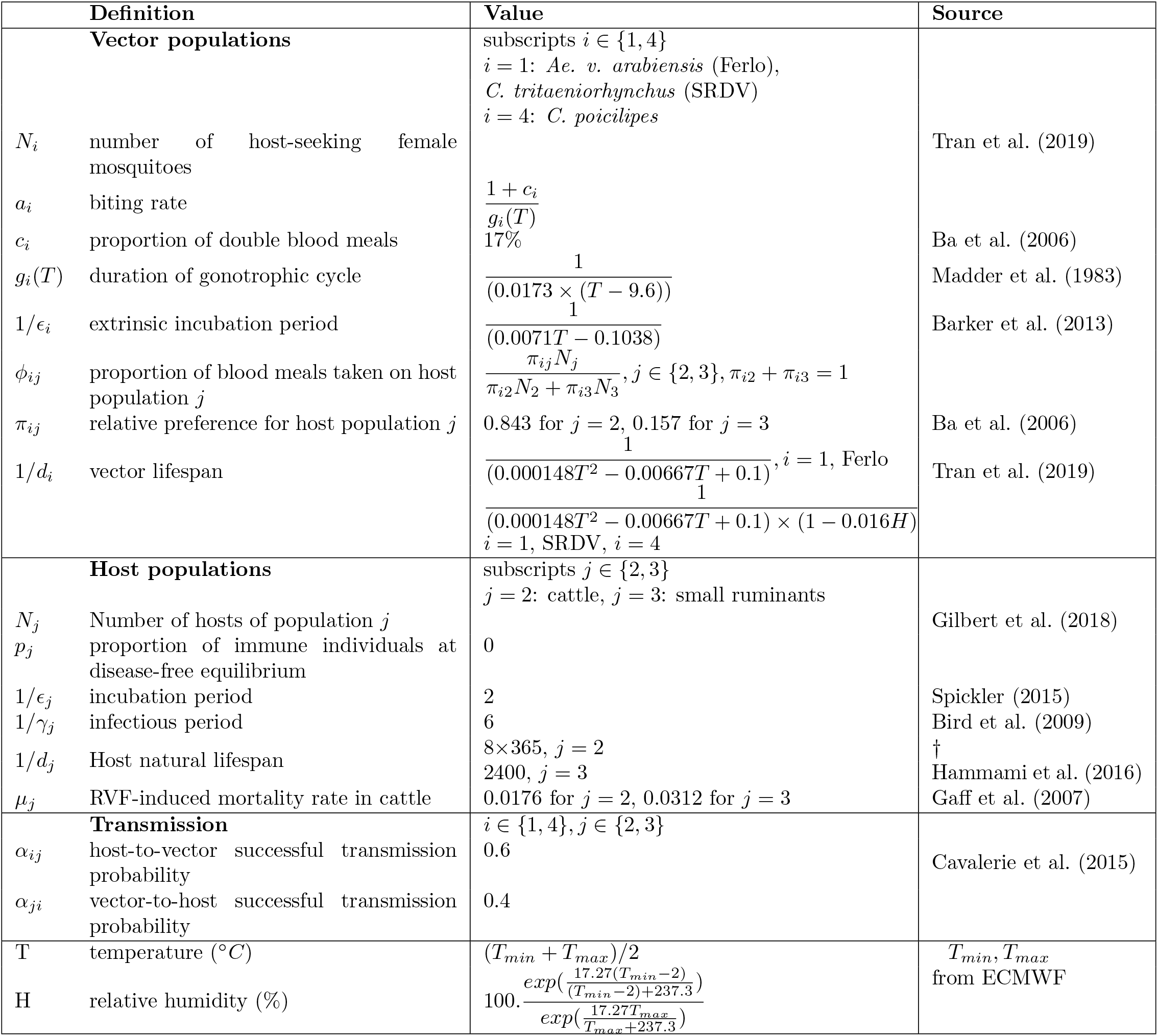
Parameter values of the basic reproduction number *R*_0_ derived from the mecanistic RVFV transmission model with two host and two vector populations. Durations are in days, rates in days^*−*1^. †: to the best of our knowledge. ECMWF : European Center for Medium Range Weather Forecasts.

### Input data

A schematic representation of the inclusion of data into our modelling framework can be found in Figure S.1.

### Spatio-temporal data on vector abundance

The vector abundance in space and time was derived from the predictions of a mechanistic model of mosquito population dynamics developed by Tran et al. (2019). This model provides the abundance of host-seeking female mosquitoes for the three vector species and the two regions of interest. Mosquito abundance is driven by rainfall, temperature, location of waterbodies, and the surface dynamics of ponds throughout the year. This model uses satellite-derived meteorological data and multispectral images to assess the habitat suitability for vectors. Tropical Applications of Meteorology using SATellite data (TAMSAT) daily rainfall estimates are used (http://www.tamsat.org.uk/cgi-bin/data/index.cgi), along with the European Centre for Medium Range Weather Forecasts (ECMWF) 10-daily minimum and maximum temperatures (https://confluence.ecmwf.int). Water bodies are detected using cloud-free Sentinel 2 scenes (level 1-C, https://earthexplorer.usgs.gov/). Their filling dynamics is estimated with an existing hydrologic model (Soti et al., 2010). The predictions of this model have been validated against entomological data collected in several sites in our study area (Biteye et al., 2018). We used weekly mosquito abundance for three consecutive rainy seasons (July to November 2014, 2015 and 2016). Our spatial units were hexagonal pixels of 1 km radius (*≃* 3.5km^2^) as in Tran et al. (2019).

### Spatial distribution of livestock

For livestock host densities, we used the Gridded Livestock of the World (GLW 3, Gilbert et al., 2018) database, which provides subnational livestock distribution data for 2010, at a spatial resolution of 0,083333^*°*^ (approximately 10km at the equator). We used the distributions of cattle and small ruminants (goats and sheeps) based on Gilbert et al. (2018) dasymetric weighting, which disaggregates census data according to weights established by statistical models using high resolution spatial covariates (land use, climate, vegetation, topography, human presence). This dataset was downscaled to match Tran et al. (2019) model pixel size by homogeneously distributing animals in smaller space units. The GLW 3 dataset is an average snapshot and does not provide time series of animal densities.

### Analytical expression of the basic reproduction number

*R*_0_ was computed only for pixels in which both hosts and vectors were present. We considered the chosen spatial resolution large enough to neglect vector dispersal among pixels, in agreement with entomological studies conducted in the Ferlo and SRDV which show that vectors rarely disperse further than 1km from ponds (Ba et al., 2005; Diallo et al., 2011; Fall et al., 2013). In addition, quantitative information on seasonal variations in livestock abundance at large scale was not available. As a result, we considered that pixels were disconnected and that animal densities remained constant.

*R*_0_ is the dominant eigenvalue of the next generation matrix of our model (Eq. 1-5). The details of its computation can be found in Supplementary Information 2. Compared to the expression derived by Turner et al. (2013) for bluetongue, we accounted for an incubation period, a natural mortality rate and a proportion of immune individuals at the disease-free equilibrium in livestock hosts. We also considered transmission probabilities (vector-to-host and host-to-vector) to be host-population specific and not only vector-population specific.

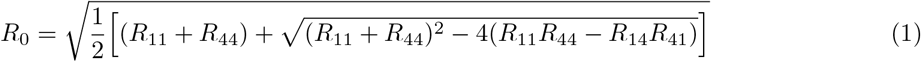

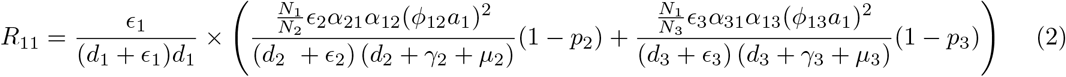

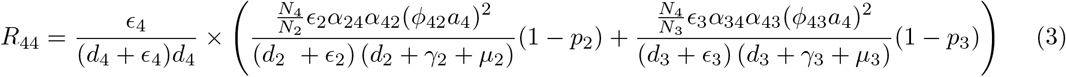

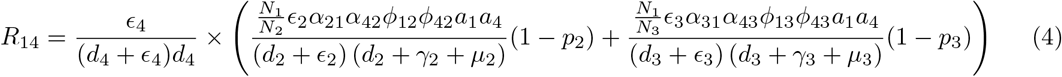

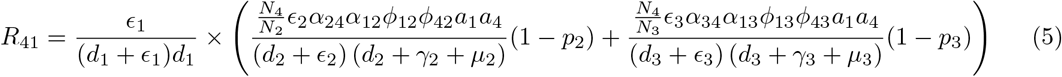

### Spatio-temporal pattern of *R*_0_

First, we identified dates and locations of RVFV introduction with high epidemic potential. For each area under study, we looked at the introduction date inducing the highest number of pixels with *R*_0_ *>* 1 each season. For clarity hereinafter, *pxl*_1_ is the number of pixels with *R*_0_ *>* 1 at a given introduction date. For each season, we computed an *R*_0_ threshold corresponding to the value of the third quartile, independently of the study area and date of introduction within a given season, *Q*_3,*year*_. We mapped pixels for which *R*_0_ *> Q*_3,*year*_ at least once within the season; we also recorded the number of times (i.e weeks) it happened during the season. For two specific locations, namely Rosso in SRDV and Barkedji in the Ferlo, we normalized *R*_0_ values (dividing them by the maximum *R*_0_ value of the season) and compared seasonnal patterns.

We then investigated the role of vector and host populations on the epidemic potential. In the Ferlo, in 2015, we looked at temporal variations of the relative abundances of vector populations within pixels with *R*_0_ *>* 1. In Barkedji, we assessed how decreased vector densities would affect the value of *R*_0_ for three different dates of virus introduction. The first date we chose corresponded to the maximum *pxl*_1_ in the Ferlo over the season. The other two dates both induced *R*_0_ *>* 1 in Barkedji but exhibited diametrically opposed vector composition, quantified with *log*_10_(*N*_4_*/N*_1_). Similarly, we looked at the effect of herd immunity, which could be acquired either through vaccination or previous exposure to RVF, on the number of pixels with *R*_0_ *> Q*_3,*year*_, per study area and season.

Finally, we performed a variance-based global sensitivity analysis using a Fourier Amplitude Sensitivity Testing (FAST, Saltelli et al., 2008). This method was used to quantify first order effects of parameters but also interactions between parameters varying simultaneously, which is not possible with “one-at-a-time” sensitivity analyses (Saltelli et al., 2019). Parameters varied within a 10% range using scaling factors (reference value of 1). A given set (scenario) of scaling factors was applied to all *R*_0_ computations of a given study area and rainy season, to maintain the spatial heterogeneity as well as the relative temporal dynamics of vector densities and temperature-dependent parameters. Temperature-dependent function formulas were kept, and temperature was not varied. We sampled 10,000 values per parameter. We tested whether introduction dates and locations with high epidemic potential were robust to these parameter variations.

## Results

Overall, there are 2.5 times more *R*_0_ values computed in the Senegal river delta and valley (SRDV) than in the Ferlo. Initially, input data provides 4419 independent pixels (1702 in the Ferlo, 2717 in SRDV) containing both hosts and vectors at least once in the 63 introduction dates (21 weeks each season) studied. Over the 236,430 possible *R*_0_ computations, 3.7% are discarded because the local vector-to-host ratio is too high (Table S.1-S.6). This mainly affects SRDV, where 52 pixels are entirely removed from the study because their ratio never goes below the chosen threshold during the 3 rainy seasons. We end up with a stable number of pixels where *R*_0_ is computed per timestep in SRDV (min 2401, max 2657 over the three seasons), whereas this number largely varies within a season in the Ferlo, minimums being 54, 7, and 5, and maximums 1673, 1702, and 1614, for 2014, 2015, and 2016 respectively. In addition, the number of pixels with *R*_0_ *>* 1 per introduction date (*pxl*_1_) never goes below 1801 for any date of introduction in SRDV (always *>*68% of *R*_0_ computations in this study area), and reaches its absolute maximum on 2016-09-12 (n = 2504). In the Ferlo, *pxl*_1_ can go from 0 for introductions in November (2014-11-24, 2015-11-30 and 2016-11-28) to 1527 (93% of *R*_0_ computations) on 2015-09-21.

In both study areas, each season, dates of introduction resulting in the highest *pxl*_1_ happen most of the time (5/6) in September (Figure 2). Seasons 2015 and 2016 exhibit similar temporal patterns of *R*_0_, both at the regional level (Figure 2A,C) and in two particular locations (Figure 2B,D). However the observed trend in 2014 is different, with an earlier peak (in August) in both regions. In addition, a third peak is observed in SRDV in November 2014. The pixel closest to Rosso has its *R*_0_ *>* 1 for every possible date of introduction over the three consecutive rainy seasons, which is not the case for the pixel closest to Barkedji.

**Figure 2:**
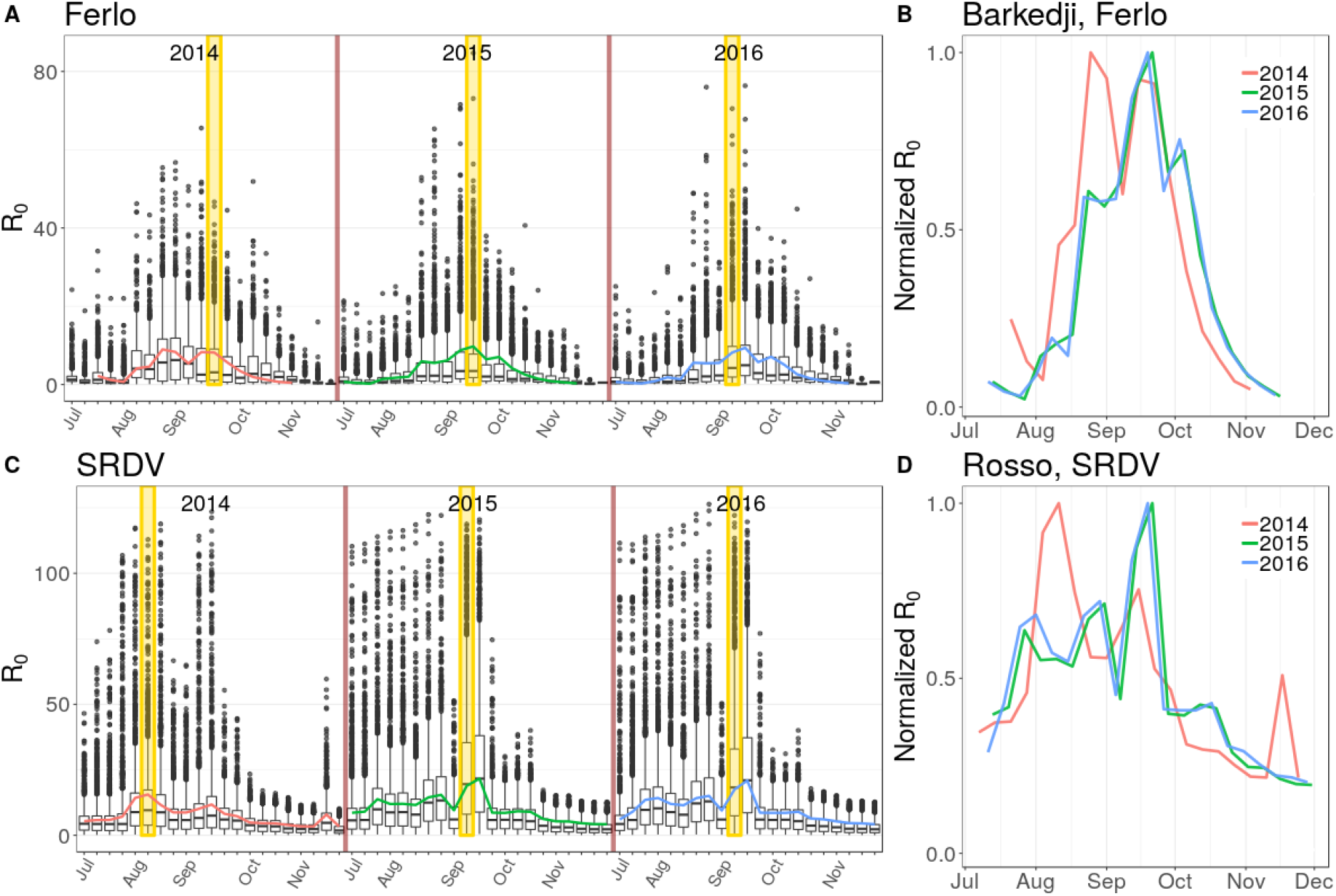
September is the month when RVF virus introduction could be the most damaging. A, C: *R*_0_ distribution by introduction week for 3 consecutive rainy seasons, spatially aggregated by region (A : Ferlo, C: SRDV). *R*_0_ values are computed independently for each introduction week, assuming constant parameters over the course of the secondary cases generation. Coloured lines show the temporal patterns of Barkedji (Ferlo) and Rosso (SRDV). Yellow bands highlight the introduction weeks inducing the highest number of pixels with *R*_0_ *>* 1, for each rainy season. Ferlo : 2014-09-22, n = 1313, 2015-09-21, n = 1527, 2016-09-12, n = 1023. SRDV, 2014-08-11, n = 2352, 2015-09-14, n = 2482, 2016-09-12, n = 2504. In the box plots, the boundaries of the box indicate the 25^*th*^ (bottom) and 75^*th*^ (top) percentile. The line within the box marks the median. Whiskers above and below the box indicate the 10^*th*^ and 90^*th*^ percentiles. Points above and below the whiskers indicate outliers outside the 10^*th*^ and 90^*th*^ percentiles. B, D: Comparison of yearly *R*_0_ pattern for Barkedji and Rosso. Values are normalized by the maximum of each rainy season.

The map of areas with highest epidemic potential varies across the three rainy seasons of 2014, 2015 and 2016 (Figure 3). In the Ferlo, the south-west of Barkedji exhibits a high epidemic potential in 2014 but not in 2015-2016. Conversely, the north-east of Barkedji exhibits a high epidemic potential in 2015-2016 but not in 2014. In SRDV, around Matam, there is a strong density of pixels with *R*_0_ above the third quartile of the season (*Q*_3,*year*_) in 2015-2016 but less so in 2014. Overall, SRDV accounts for a larger proportion of pixels with *R*_0_ *> Q*_3,*year*_ than the Ferlo, every season (at least three times more, Table S.7). In addition, *R*_0_ values above *Q*_3,*year*_ appear less often per pixel in the Ferlo than in SRDV, every season (Figure 3, pixels ranging from green to yellow, Table S.7).

**Figure 3:**
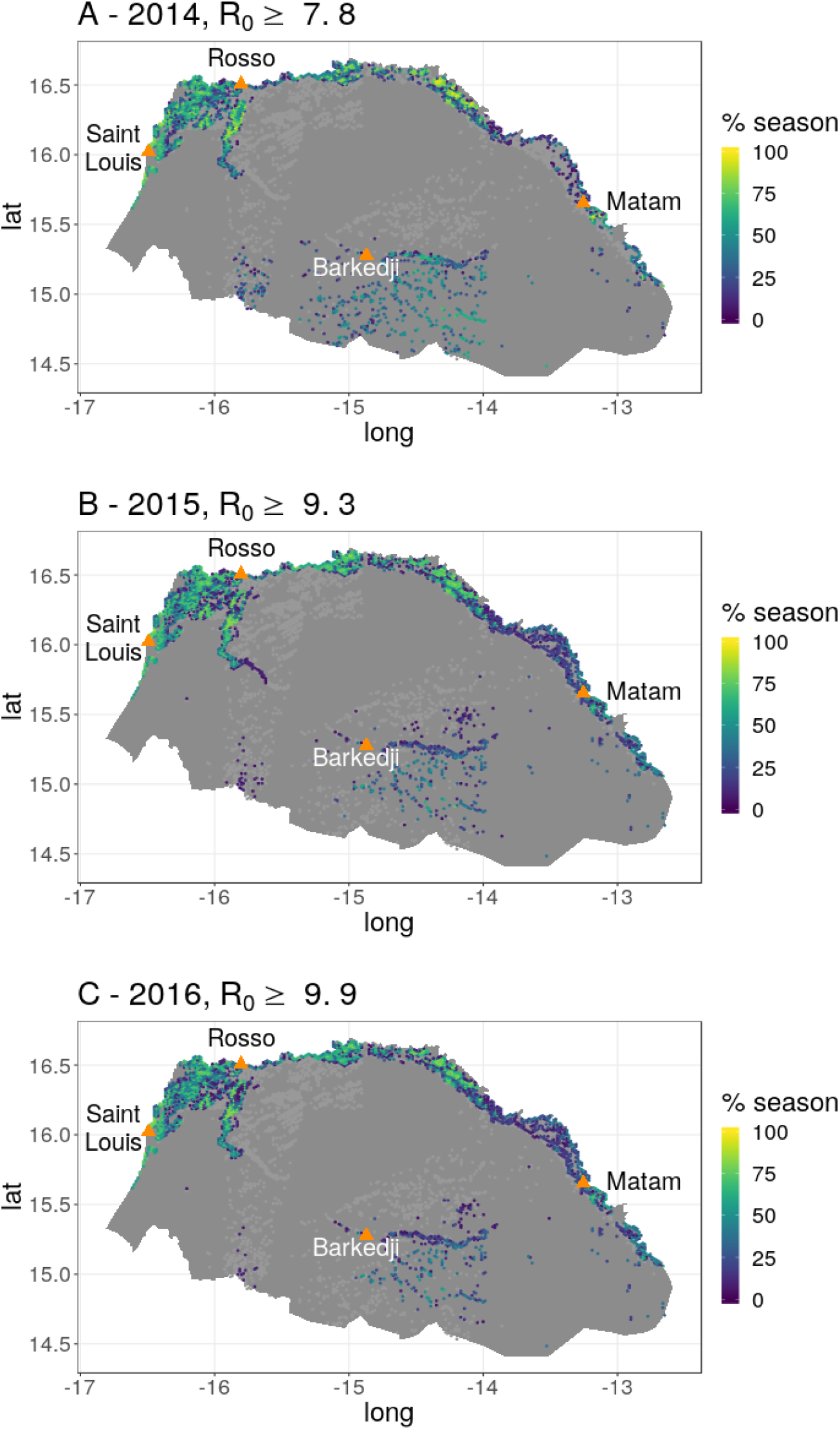
Zones of high RVF epidemic potential change between rainy seasons. Map of northern Senegal showing pixels with *R*_0_ *≥ Q*_3,*year*_ (third quartile of *R*_0_ values) at least once in the season. Pixels are coloured by percentage of season spent above the threshold (1 to 21 weeks). Orange points are important locations to ease figure reading. Lights grey pixels are other pixels where *R*_0_ is computed during the season.

The results of the sensitivity analysis show that in both study areas and for each season, dates and locations of RVFV introduction resulting in high epidemic potential are robust to parameter variations. Dates of introduction inducing the highest *pxl*_1_ are similar between scenarios (Table S.8), as well as the distribution of pixels with high *R*_0_ values (*R*_0_ *> Q*_3,*year*_) and the number of times for which it happens for those pixels (Figure S.8).

In the Ferlo, *Ae. v. arabiensis* tends to be the most abundant vector population within pixels with *R*_0_ *>* 1 at the beginning of the rainy season, while *C. poicilipes* is the most abundant later in the season (Figure 4A, Figure S.4). Nonetheless, the vector composition shows a large variability between pixels for a same date of introduction. For instance on October 12^*th*^ 2015, minimum and maximum relative abundances are *log*_10_(*N*_4_*/N*_1_) = *−*3.74 and *log*_10_(*N*_4_*/N*_1_) = 4.44 respectively (Figure 4A). In addition, when looking at dates resulting in the highest *pxl*_1_, *Ae. v. arabiensis* is on average the most abundant in pixels with *R*_0_ *>* 1 in 2014 (2014-09-22, Figure S.4), while *C. poicilipes* is on average the most abundant in pixels with *R*_0_ *>* 1 in 2015 and 2016 (2015-09-21, 2016-09-12, Figure 4A, Figure S.4). In Barkedji, diametrically opposed vector compositions can induce *R*_0_ *>* 1, such as 2015-08-24 when *log*_10_(*N*_4_*/N*_1_) = *−*1.08 and 2015-10-12 when *log*_10_(*N*_4_*/N*_1_) = 1.08 (Figure 4A,B,D). In SRDV, *C. tritaeniorhynchus* is always the most abundant species in every pixel with *R*_0_ *>* 1, but the difference between the two populations is less important than in the Ferlo (Figure S.4).

**Figure 4:**
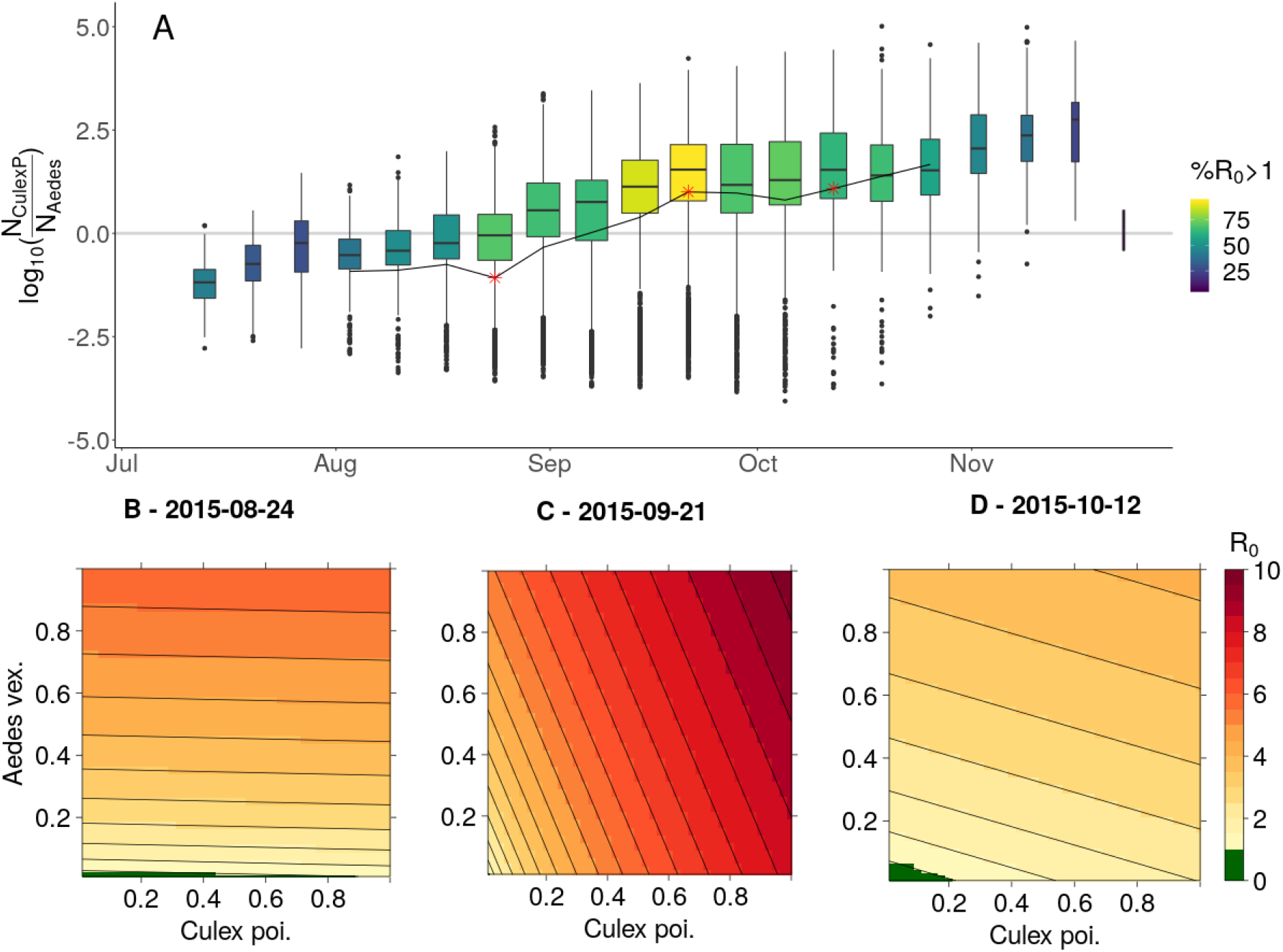
Decreased vector densities do not highly reduce RVF epidemic potential in at-risk locations. Example of Ferlo 2015. A: Relative abundance of vector populations *log*_10_(*N*_4_*/N*_1_) within pixels having *R*_0_ *>* 1 over time. Light grey line indicates equal densities. For boxplots, see legend of Figure 2. Boxplots width is proportionnal to the number of pixels with *R*_0_ *>* 1 (*pxl*_1_, min 4, max 1527). Colours inside box plots indicate the proportion of pixels with *R*_0_ *>* 1 among those where *R*_0_ is computed (min 7%, max 93%). Black line corresponds to the particular value of the ratio in Barkedji, for introduction weeks inducing *R*_0_ *>* 1. Stars are positionned at introduction weeks 2015-08-24, 2015- 09-21 and 2015-10-12. 2015-09-21 corresponds to the maximum *pxl*_1_ in the Ferlo this season. The other two dates both induce *R*_0_ *>* 1 in Barkedji but exhibit diametrically opposed vector composition. B-D: Variation of *R*_0_ in Barkedji when decreasing vector densities, for 3 different weeks of introduction. Axes represent the proportion of initial vector density applied for the *R*_0_ computation, the reference is at the top right corner (1, 1).

Decreased vector densities can hardly reduce *R*_0_ values of at-risk pixels below 1 (Figure 4). In Barkedji, this is observed regardless of RVFV introduction date and whichever species is the most abundant. In addition, the vector composition is not an indicator of which population, if decreased, will more strongly affect *R*_0_. Indeed, for RVFV introductions on August 24^*th*^ 2015 and September 21^*st*^ 2015, decreasing the density of the most abundant vector population has the most important effect on *R*_0_ value (Figure 4A,B,C). This is not observed on October 12th 2015, when *C. poicilipes* are more numerous than *Ae. v. arabiensis* in Barkedji (Figure 4A, third red star), but decreasing the density of *Ae. v. arabiensis* has the strongest impact on *R*_0_ (Figure 4D).

In both study areas, an increase in the proportion of immune cattle decreases the number of pixels with high *R*_0_ values (*R*_0_ *> Q*_3,*year*_) more effectively than increasing the proportion of immune small ruminants (Figure 5B,C, Figure S.5). In most pixels (4302/4367 = 98,5%), the number of small ruminants is higher than the number of cattle (Figure 5A). However, the difference in host populations sizes is smaller in SRDV than in the Ferlo. Indeed, there are on average 7.5 times more small ruminants than cattle in the Ferlo, and only twice more in SRDV. This is related to the presence of both very low cattle densities and very high small ruminant densities in the Ferlo, while the range of SRDV host distributions is narrower (Figure S.3). As a consequence, since cattle are in fewer numbers than small ruminants while being the prefered host of all vector species studied. (Table 1, Supplementary information 1.2) they are more likely to get bitten more than once. These bites, provided they result in successful transmission (first to the host, then to a vector), can strongly contribute to RVF epidemic potential.

**Figure 5:**
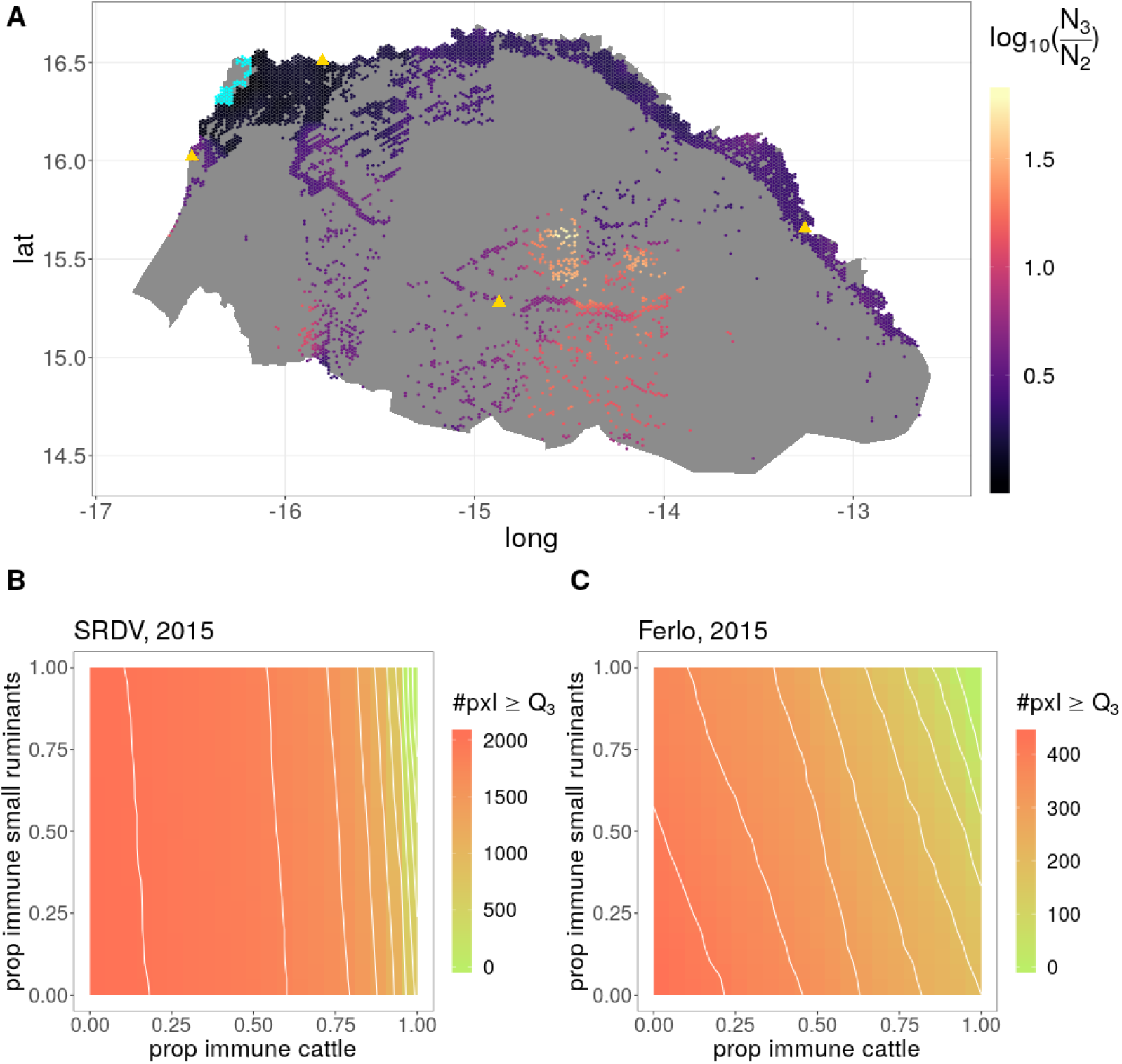
In both study areas, an increase in the proportion of immune cattle decreases the number of pixels with high *R*_0_ values (*R*_0_ *> Q*_3,*year*_) more effectively than increasing the proportion of immune small ruminants. A - Map of relative densities of hosts *log*_10_(*N*_3_*/N*_2_) within pixels of our study area. Blue pixels have more cattle than small ruminants, the biggest difference being *log*_10_(*N*_3_*/N*_2_) = *−* 0.08. B-C - Variation of the number of pixels with *R*_0_ *> Q*_3,2015_ by study area (B : SRDV, C : Ferlo) when increasing host immunity. Axis represent proportion of immune hosts applied for the *R*_0_ computation. The reference is the absence of herd immunity, (0, 0), in the bottom left corner.

Finally, in the Ferlo, every rainy season, the feeding preferences and the gonotrophic cycle duration of the most abundant vector species are the most influential parameters on the epidemic potential at the region scale (Figure 6, Figure S.6). In 2015, the first order effects of these parameters explain respectively 47% and 19% of the variance observed in *pxl*_1_, for the date of introduction inducing the highest *pxl*_1_ (Table S.8). In SRDV, *pxl*_1_ does not vary much in our sensitivity analysis (maximum 3% from the reference value in 2016, Figure S.7) for the dates of introduction inducing the highest *pxl*_1_, because *pxl*_1_ quickly reaches the total number of pixels where *R*_0_ is computed for the study area. It is nonetheless influenced by the same parameters than highlighted for the Ferlo (Figure S.7). Precisely, the more the feeding preference of the most abundant vector population is skewed towards cattle, and the more often these vectors have to take a new blood meal, the higher *pxl*_1_.

**Figure 6:**
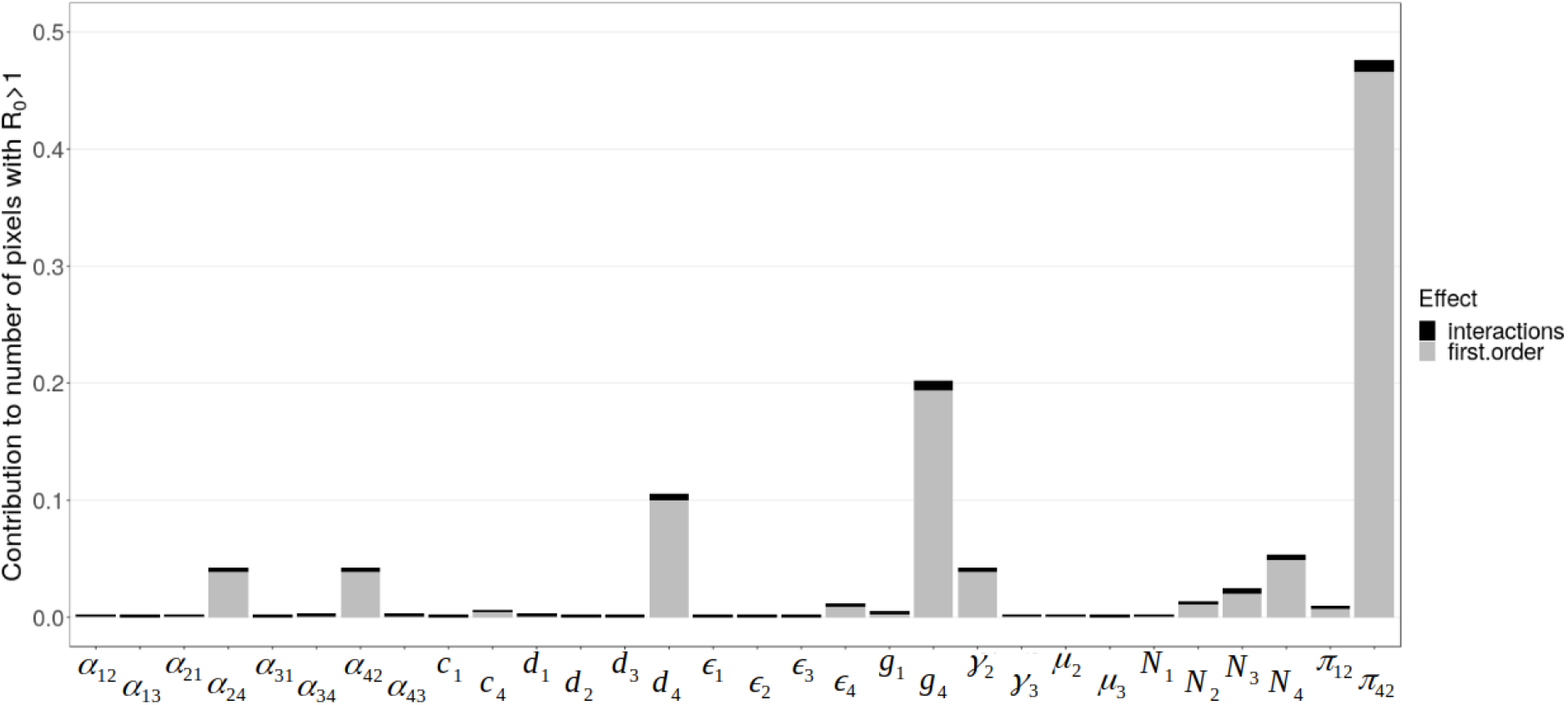
The feeding preferences and the gonotrophic cycle duration of the most abundant vector species are the most influential parameters on the epidemic potential at the region-scale. Example of Ferlo, 2015. Results of the FAST sensitivity analysis showing contribution of model parameters to the number of pixels with *R*_0_ *>* 1, *pxl*_1_, for the introduction week inducing the highest *pxl*_1_ of the rainy season. Sensitivity indices for principal effect in grey and for interactions in black. Definition and reference values of parameters can be found in Table 1. The introduction week inducing the highest *pxl*_1_ during the 2015 rainy season is 09-21 for 299,999 scenari and 09-14 for one scenario (Table S.8). At these dates, *C. poicilipes* is the most abundant vector population in the Ferlo (Figure S.4)

## Discussion

The results of our analyses show that an introduction of Rift Valley fever virus in September has the potential to trigger an epidemic almost everywhere in northern Senegal. Areas with high epidemic potential during most of the rainy season also exists, particularly in the Senegal river delta and valley, due to the continuous presence of water. In contrast, in the Ferlo region, the most at-risk ponds change between rainy seasons. These results are robust to parameter variations tested in our sensitivity analysis, following a global variance-based approach requested for models with nonlinearities and interactions (Saltelli et al., 2019).

We provide the first mapping of RVF epidemic potential in the West African Sahel using the basic reproduction number. We achieve better spatial and temporal resolutions than previous studies in RVF-free regions (Barker et al., 2013; Fischer et al., 2013), which was made possible by the use of satellite Sentinel 2 images by Tran et al. (2019) along with ground truth validation data and a precise knowledge of temporary ponds filling dynamics. However, host densities, which do not stand out in our sensitivity analysis, may vary beyond the range presentely allowed. Indeed, their temporal dynamics, mostly driven by animal mobility, is not incorporated into GLW 3 data, and might affect the population-specific contact rate with vectors and therefore *R*_0_ values. Remote-sensing methods are considered a promising tool to measure human mobility (Bharti and Tatem, 2018), but we also need qualitative data on factors guiding decisions of nomadic herders in order to include animal mobility in a mechanistic way (Apolloni et al., 2018; Belkhiria et al., 2019).

The mechanistic approach used in this paper is the best suited to describe the complexity of RVF epidemic potential in our study region. Indeed, neither host nor vector densities alone are sufficient to predict the local epidemic potential, contrary to what was implied by previous mappings and statistical studies (Bicout and Sabatier, 2004; Caminade et al., 2011). Indeed, it is actually the process of blood feeding, during which host and vector populations interact, which should accurately be described to achieve the most reliable estimates of RVF epidemic potential. We account for the influence of temperature in our model, which is known to strongly influence the risk of vector-borne diseases transmission (Mordecai et al., 2017; Kamiya et al., 2020; Mordecai et al., 2019), but we do not simulate the consequences of global changes, which is beyond the scope of this study. The adequate contact rate, an aggregated parameter used in previous models (Gaff et al., 2007; Mpeshe et al., 2014), is decomposed here to assess the importance of each of its components, as in Turner et al. 2013. Based on our sensitivity analysis, we recommend that future biological investigations focus on the feeding preferences of vectors and the duration of their gonotrophic cycle, in relation with temperature.

The inclusion of two host and two vector populations provides new insights on RVF epidemic potential, and this structure should be kept for future models studying RVF in the region. We show that decreased vector densities are not sufficient to limit the epidemic potential of RVF, regardless of the introduction date considered. Indeed, vector abundances are not always a good predictor of RVF epidemic potential, with high *R*_0_ values sometimes driven by the least abundant vector population. Moreover, cattle contribute strongly to RVF transmission and their immune status is likely to influence the epidemic potential at the regional scale. Favoring vaccination of cattle over small ruminants is not what is usually done in the field. Veterinary services, along with herders, tend to promote small ruminant vaccination as they are more likely to die from the disease and thus need more protection (Sow et al., 2016). The importance of small ruminant trade, particularly around the Tabaski festival, might also justify this approach (Lancelot et al., 2019). Operationnal decisions regarding targeted vaccination campaigns should therefore consider the potential benefits of cattle immunity at the population scale.

In the present study, we provide a better understanding of conditions which could trigger the onset of an epidemic. However, this should be interpreted with caution, and should not be considered as an indicator of epidemic size. Indeed, multiple introductions or sudden unfavourable conditions might lead to diseases persisting with *R*_0_ *<* 1 or dying out with *R*_0_ *>* 1 (Li et al., 2011). In addition, our results could be used as initial conditions for a stochastic mecanistic model of spatio-temporal transmission, which would include processes underlying epidemic dynamic after RVFV introduction, such as animal mobility. Such a model would benefit from an increased availability of epidemiological data for validation and parametrisation, which are necessary to unravel the underlying mechanisms driving the spatio-temporal dynamics of RVF in the West African Sahel.

### Data and code accessibility

Input data and scripts are available online: http://sourcesup.renater.fr/www/rvf-r0-senegal/

## Data Availability

Input data and scripts are available online

http://sourcesup.renater.fr/www/rvf-r0-senegal/

## Author contributions

**HC**: Conceptualization, Data curation, Formal analysis, Funding acquisition, Methodology, Software, Validation, Visualization, Writing - original draft, Writing - review & editing. **RM**: Conceptualization, Methodology, Supervision, Visualization, Writing - original draft, Writing - review & editing. **AGF**: Resources, Writing - review & editing. **MML**: Resources, Writing - review & editing. **RL**: Conceptualization, Funding acquisition, Project administration, Supervision, Writing - review & editing. **PE**: Conceptualization, Funding acquisition, Methodology, Project administration, Resources, Supervision, Validation, Visualization, Writing - original draft, Writing - review & editing.

